# PU-Boost: A Two-Stage Reliable-Sample Reconstruction Positive-Unlabeled Learning Framework for Screening Undiagnosed Hypertension

**DOI:** 10.64898/2026.10.04.26364708

**Authors:** Huanqi Wu, Liangkun Shi, Hanfei Yang, Boyang Li, Qiuyu Wang, Zhuofan Zhan, Lizhu Guo

## Abstract

In hypertension screening, previously diagnosed cases provide reliable positive labels, whereas individuals without a prior diagnosis comprise both undiagnosed hypertensive and non-hypertensive individuals, creating a canonical positive-unlabeled (PU) learning setting. We propose PU-Boost, a two-stage reliable-sample reconstruction framework that uses only readily obtainable demographic, behavioral, medical-history, and anthropometric variables. During training, only previously diagnosed hypertension cases are treated as labeled positives (P), while all remaining individuals are treated as unlabeled (U); no additional reference labels are supplied to the two-stage reconstruction procedure. Elastic Net first provides a global ranking of P versus U, after which a random forest refines the uncertain region and highly ambiguous observations are rejected. XGBoost is then trained on the reconstructed class-balanced pseudo-labeled sample. In the independent test set, PU-Boost achieved a sensitivity of 0.817 and a balanced accuracy of 0.682, the highest observed values among the compared models. It produced 729 false negatives, 41.1% fewer than standard XGBoost, although specificity decreased to 0.547. SHAP analysis indicated that established hypertension-related features, including age, body mass index, waist circumference, and family history, contributed strongly to model output. PU-Boost therefore provides a statistically motivated framework for reducing missed cases when screening for undiagnosed hypertension under incomplete diagnostic labeling.

## 1 Introduction

Hypertension is a major global public-health challenge and an important risk factor for cardiovascular disease, stroke, chronic kidney disease, and premature mortality(Collaboration, 2021; Oparil et al., 2018; World Health Organization, 2025). Although hypertension can be identified directly through blood-pressure measurement, a substantial proportion of affected individuals remain undiagnosed in real-world settings, particularly during earlier stages when symptoms are often absent(World Health Organization, 2025). National hypertension surveys in China indicate that prevalence remains high while awareness, treatment, and control remain suboptimal(Cao et al., 2025; Wang et al., 2018). Undiagnosed hypertension may delay timely intervention and increase the long-term burden on health-care systems. Accordingly, identifying high-risk individuals using simple and readily available information is an important objective in hypertension prevention and management.

Machine-learning methods have increasingly been used for hypertension risk prediction and disease screening. Previous studies have developed predictive models from demographic characteristics, lifestyle factors, anthropometric measures, and electronic health records using logistic regression, random forests, gradient boosting, and neural networks(Chowdhury et al., 2022; Silva et al., 2022). However, when training labels are constructed solely from prior diagnosis, conventional supervised classifiers typically treat previously diagnosed individuals as positives and everyone else as non-positive(Chang et al., 2022). If hidden positives are present among those without a prior diagnosis, the observable label S differs from the reference disease status Y(Chang et al., 2022; Elkan & Noto, 2008). Consequently, minimizing empirical loss with respect to S targets a quantity closer to Pr(S=1|X) than to the screening target Pr(Y=1|X). This target mismatch causes undiagnosed reference-positive individuals to contribute negative supervision and can distort the learned decision boundary. Based on prior diagnosis and reference disease status, participants can be classified retrospectively into three groups: group A, diagnosed hypertension; group B, undiagnosed hypertension; and group C, non-hypertensive individuals. The A/B/C grouping is used to describe the reference label structure, to support model selection in the validation set, and to evaluate final performance. During the two-stage PU reconstruction itself, only group A is observed as a reliable positive class, whereas the reference identities of groups B and C are hidden and jointly form the unlabeled population. Direct blood-pressure measurements are used only to construct Y and for downstream validation and evaluation, and are not included among the predictors. Thus, hidden reference-positive and reference-negative individuals may overlap extensively in the available feature space. The central modeling problem is therefore not simply to increase classifier complexity, but to identify high-confidence observations within the unlabeled population while progressively restricting the region of label uncertainty without accessing B/C reference identities.

**Figure 1.**
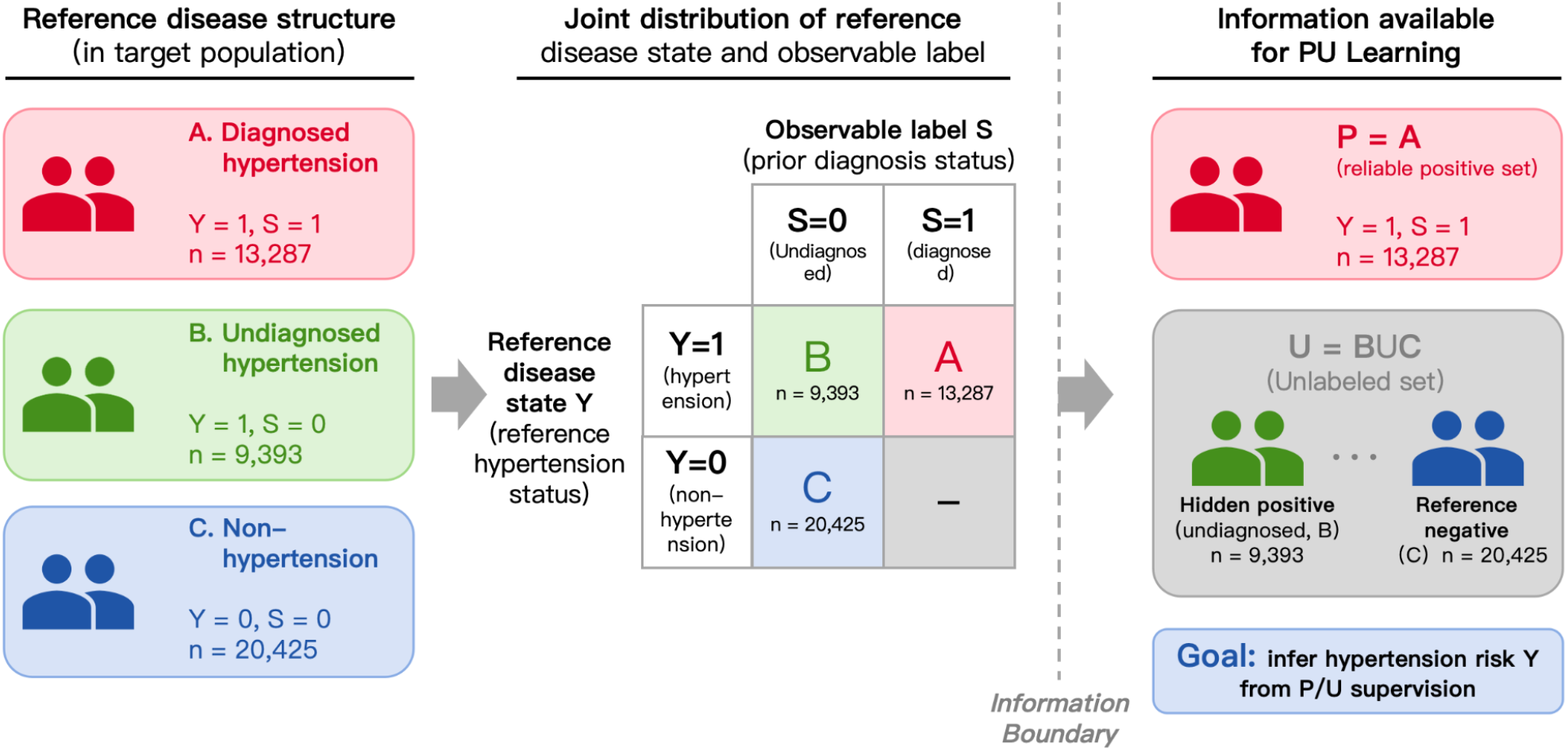
Reference disease structure and information available during positive-unlabeled learning. Participants were retrospectively classified as diagnosed hypertension (group A; Y=1, S=1), undiagnosed hypertension (group B; Y=1, S=0), and non-hypertensive (group C; Y=0, S=0). During two-stage PU reconstruction, only group A was observed as the labeled-positive set P, whereas groups B and C were indistinguishable to the training algorithm and jointly formed the unlabeled set U. Reference B/C identities were used only outside the reconstruction procedure for model selection and performance evaluation.

Positive-unlabeled (PU) learning addresses classification problems in which reliable positive examples are available but the remaining observations are unlabeled(Bekker & Davis, 2020; du Plessis et al., 2014; Elkan & Noto, 2008). Let P denote the labeled-positive set and U the unlabeled set. Samples in P carry an observed positive label, whereas the latent class membership of U is unknown and U contains both hidden positives and negatives. The training-information structure in this study follows this setting: *P* = *A* and *U* = *B* ∪ *C*, with B/C reference identities unavailable to the training algorithm. Screening for undiagnosed hypertension can therefore be naturally formulated as a PU-learning problem. Unlike conventional supervised learning that treats U as a negative class, PU learning seeks to exploit information in P and the mixed unlabeled set U to construct a decision rule informative about the reference disease state(Bekker & Davis, 2020; du Plessis et al., 2015; Kiryo et al., 2017).

PU learning has been applied to cancer prediction, Alzheimer’s disease identification, and other disease-screening tasks, but its use for screening undiagnosed hypertension using only readily available demographic, lifestyle, medical-history, and anthropometric variables remains limited(Li et al., 2022; Tran et al., 2025; Wu et al., 2023; Zhou et al., 2022). In the present setting, the challenge arises not only from the absence of explicit negative labels but also from substantial overlap within U. A single-stage procedure that assigns pseudo-labels to all unlabeled observations may force highly uncertain samples near the decision boundary into either class and propagate pseudo-label error to the downstream classifier(Bekker & Davis, 2020; Hendrickx et al., 2024; Liu et al., 2002). We therefore focus on progressively identifying high-confidence candidate samples from U while allowing persistently ambiguous observations to remain unassigned and limiting their contribution to the final supervised loss.

Using data from 43,105 community-dwelling adults aged ≥45 years across eight study areas in China, we develop PU-Boost, a two-stage PU-learning framework. In stage 1, Elastic Net logistic regression produces a regularized P-versus-U discrimination score for global stratification into high-confidence candidate positives, candidate negatives, and an uncertain region. In stage 2, a random forest is applied only to the stage-1 uncertain region to exploit nonlinear structure and interactions, yielding additional candidate positives, candidate negatives, and a highly ambiguous subset that is excluded from final classifier training. The retained high-confidence candidates are then assembled into a balanced pseudo-labeled training set and used to train XGBoost as the final classifier. The architecture progressively restricts uncertain supervision through global stratification, local nonlinear refinement, and uncertainty rejection.

This study makes three main contributions. First, it formalizes screening for undiagnosed hypertension as a PU-learning problem and explicitly distinguishes the observable diagnosis label from the reference disease status. Second, it introduces a two-stage reliable-sample reconstruction strategy combining global stratification, local nonlinear refinement, and rejection of persistently uncertain observations, thereby avoiding forced pseudo-label assignment for the entire unlabeled set. Third, it evaluates the screening operating characteristics of the resulting model in an independent test set, with particular emphasis on sensitivity, false negatives, specificity, balanced accuracy, and model interpretability, and compares these characteristics with conventional supervised-learning baselines.

## 2 Methods

### 2.1 Study population and data source

This study used data from a nationally representative, cross-sectional, community-based survey conducted in China between 2014 and 2016(Du et al., 2021). The original survey used a two-stage stratified cluster-sampling design, with sampling sites in Jilin, Xinjiang, Beijing, Henan, Jiangxi, Zhejiang, Guangdong, and Yunnan, covering seven major geographic regions of China: Northeast, Northwest, North, Central, East, South, and Southwest. Data collection was completed for 47,841 adults aged ≥45 years, of whom 43,105 met the analytic inclusion criteria for the present study. The survey collected demographic characteristics, lifestyle and behavioral factors, medical history, medication information, blood-pressure measurements, laboratory measures, and anthropometric indices. The sampling design, measurement procedures, and data-collection protocol of the original survey have been described previously(Du et al., 2021). The study protocol was approved by the Ethics Committee of Beijing Anzhen Hospital (IRB No. 2013024), and all participants provided written informed consent.

### 2.2 Predictors and data preprocessing

The study was designed to develop a screening model for risk stratification before confirmatory blood-pressure measurement. Predictors were therefore restricted to demographic characteristics, lifestyle factors, general medical history, and anthropometric measures that are relatively easy to obtain at the population level. Medication information, laboratory measurements, and blood-pressure measurements used to define the reference hypertension status were excluded from model inputs. Following problem-oriented variable screening, 60 predictors were retained from approximately 320 original candidate variables for model development.

To prevent information leakage, all preprocessing steps requiring estimation from the data distribution were performed within the training set and then applied unchanged to the validation and test sets. Observations with missing values in variables required for model development were excluded using complete-case analysis. Continuous variables were standardized using parameters estimated from the training set. Categorical variables were represented by dummy (indicator) variables with one level designated as the reference category; category levels and encoding rules were defined from the training data and applied consistently to the validation and test sets.

### 2.3 Definition of hypertension status and label groups

The reference hypertension status was defined jointly from prior diagnosis of hypertension and standardized blood-pressure measurements obtained during the survey. Under the original survey protocol, each participant underwent three blood-pressure measurements. Let *SBP*_*im*_ and *DBP*_*im*_ denote the systolic and diastolic blood pressure, respectively, for participant *i* at measurement *m*, where *m* = 1, 2, 3. The participant-level representative blood pressures were defined as the means of the three measurements:

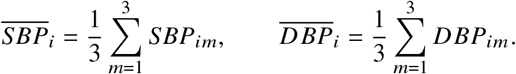

Following the hypertension criterion used in the original survey (Du et al., 2021; Revision., 2019)(systolic blood pressure 140 mmHg; diastolic blood pressure 90 mmHg), define the measured elevated-blood-pressure indicator *H*_*i*_ :

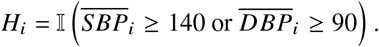

Define the prior-diagnosis indicator *Di* ∈ {0, 1}: if participant i had previously received a diagnosis of hypertension, then *D*_*i*_ = 1; otherwise *D*_*i*_ = 0. Antihypertensive medication use was not treated as an independent criterion for the reference label. Direct blood-pressure measurements were used only to construct the reference disease status and for subsequent performance evaluation and were not included as model predictors.

Based on the combination of *D*_*i*_ and *H*_*i*_, participants were retrospectively divided into three reference groups:

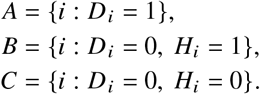

Group A comprised participants with previously diagnosed hypertension; group B comprised participants without a prior diagnosis whose measured blood pressure met the hypertension criterion; and group C comprised participants without a prior diagnosis whose measured blood pressure did not meet the hypertension criterion. In the complete analytic cohort, groups A, B, and C contained 13,287, 9,393, and 20,425 participants, respectively.

Define the reference disease status *Y*_*i*_ ∈ {0, 1}:

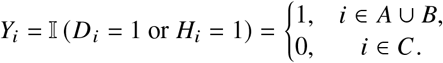

Unlike *Y*_*i*_, the observable training label *S*_*i*_ records only prior diagnosis status:

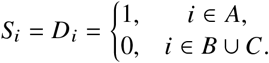

Thus, the three reference groups correspond to label structures *A* : (*Y* = 1, *S* = 1), *B* : (*Y* = 1, *S* = 0), and *C* : (*Y* = 0, *S* = 0). The reference identities of groups B and C are used only outside the two-stage reconstruction procedure, including model selection, verification, and performance evaluation, and are not provided as inputs to PU-Boost during the two-stage training process. The labeled-positive set *P* and unlabeled set *U* used for training are defined as

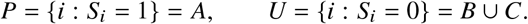

Therefore, *S*_*i*_ = 1 → *Y*_*i*_ = 1, whereas *S*_*i*_ = 0 contains both reference-positive and reference-negative individuals.

**Figure 2.**
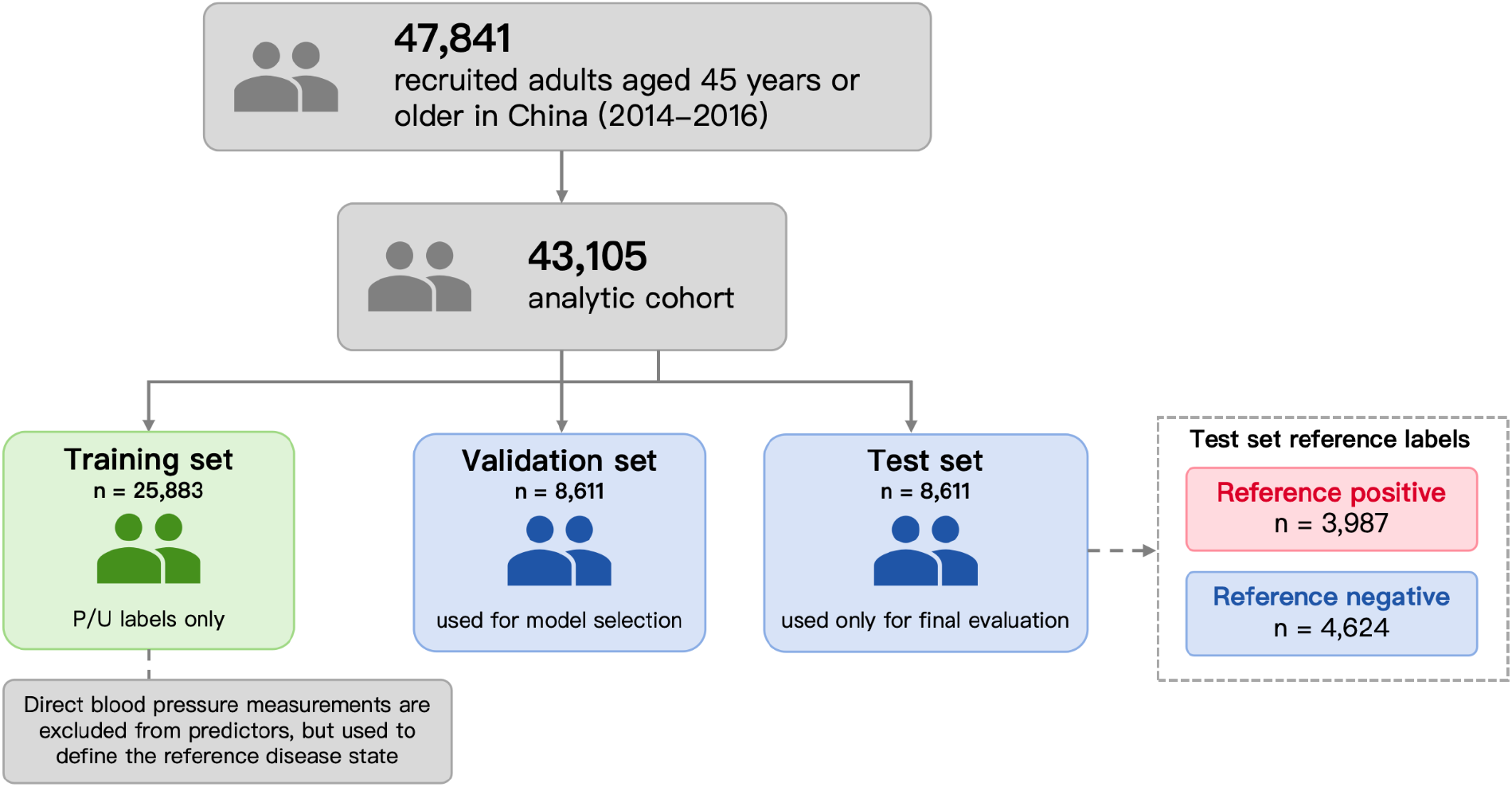
Cohort construction, data partitioning, and information isolation. Data collection was completed for 47,841 adults aged ≥45 years, Participants with missing information in variables required for model development were excluded, yielding a final analytic sample of 43,105. Participants were divided into training (n=25,883), validation (n=8,611), and test (n=8,611) sets in a 6:2:2 ratio. Two-stage PU reconstruction used only P/U information from the training set; reference disease status Y was used for validation-based model selection and final test-set evaluation. Direct blood-pressure measurements were excluded from model predictors.

### 2.4 Dataset partitioning and information isolation

The 43,105 participants were randomly divided in a 6:2:2 ratio into training, validation, and test sets containing 25,883, 8,611, and 8,611 participants, respectively. Two-stage PU reconstruction was performed only in the training set, and the training algorithm accessed only the *S*-defined *P*/*U* information. The reference disease status *Y* in the validation set was used only for model selection and hyperparameter comparison and did not contribute to training-sample reconstruction; the test set was used only for final performance evaluation after model specification was fixed. This information separation prevented B/C reference identities from entering the two-stage PU-Boost fitting procedure.

### 2.5 Formalization of the positive-unlabeled learning problem

If the observable label *S* is used directly as the supervised response, the classifier targets a quantity closer to Pr(*S* = 1 | *X*), whereas screening for undiagnosed hypertension is concerned with Pr(*Y* = 1 | *X*). Under the reliable-positive assumption *S* = 1 → *Y* = 1,

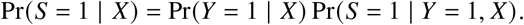

Let c(X)=Pr(S=1|Y=1,X), so that Pr(S=1|X)=c(X)Pr(Y=1|X). If c(X) is constant, the two probabilities remain proportional but are generally not equal; if c(X) varies with X, modeling the observable diagnosis label may also distort the ordering of individual disease risk(Jain et al., 2016; Mielniczuk & Wawrzeńczyk, 2025). PU-Boost therefore does not interpret the stage-1 P-versus-U output as an absolute disease probability. Instead, it uses *P* and *U* to construct a continuous screening score and reconstructs the final supervised training set through selective pseudo-labeling and rejection of uncertain observations.

### 2.6 Assessment of distributional overlap within the unlabeled set

To characterize overlap between reference-positive group B and reference-negative group C within the unlabeled set U, exploratory between-group analyses and low-dimensional visualizations were conducted outside the model-training procedure using the reference labels. Differences in the means and variances of continuous variables were examined descriptively using two-sample t tests and F tests, respectively. These analyses were used only to characterize task difficulty and individual-level separability; they were not used to fit either stage of the PU model or to construct training thresholds.

### 2.7 Two-stage reliable sample reconstruction

PU-Boost applies a two-stage selective pseudo-labeling procedure under the P/U information structure. In stage 1, a regularized linear model provides a global P-versus-U ranking and partitions the training sample into high-confidence candidate positives, candidate negatives, and a stage-1 uncertain set.

Stage 2 refines only the uncertain set using a nonlinear model, while observations that remain insufficiently certain after both stages are assigned to a reject set. Rejected observations are excluded from final binary supervised training, thereby avoiding forced pseudo-label assignment for highly ambiguous samples.

#### Stage 1: Global P-versus-U ranking with Elastic Net

Let *X*_*i*_ ∈ R^*P*^ denote the feature vector for participant *i*. Stage 1 uses the observable training label *S*_*i*_ as the response, coding observations in *P* as 1 and observations in *U* as 0. Elastic Net logistic regression produces the stage-1 P-versus-U score

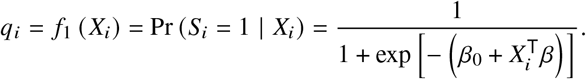

*q*_*i*_ measures the extent to which an observation resembles the labeled-positive set and is used for ranking and stratification(Friedman et al., 2010; Zou & Hastie, 2005); it is not interpreted as a calibrated estimate of Pr (*Y*_*i*_ = 1 | *X*_*i*_). Parameters are estimated by minimizing the penalized negative log-likelihood:

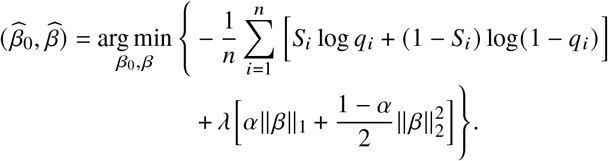

Where

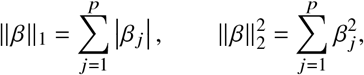

λ controls the overall strength of regularization, and α controls the relative contribution of the L1 and L2 penalties; α = 1 corresponds to LASSO, α = 0 corresponds to Ridge regression, and 0 < α < 1 combines both penalties. The pair (α, λ) is selected by 10-fold cross-validation. If L_*K*_ (α, λ) denotes the validation loss in fold *K*, then

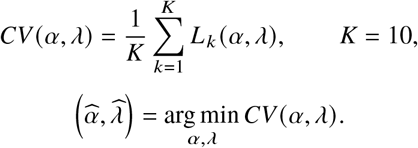

Let the stage-1 high- and low-confidence thresholds be 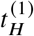 and 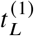, respectively, with 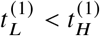 . The stage-1 partition is

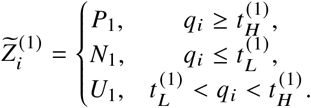

Here, *P*_1_, *N*_1_, and *U*_1_ denote the stage-1 candidate-positive set, candidate-negative set, and uncertain set, respectively. All set memberships are determined solely from P/U information available during training and from training-internal threshold rules; they do not represent observed reference truth.

**Figure 3.**
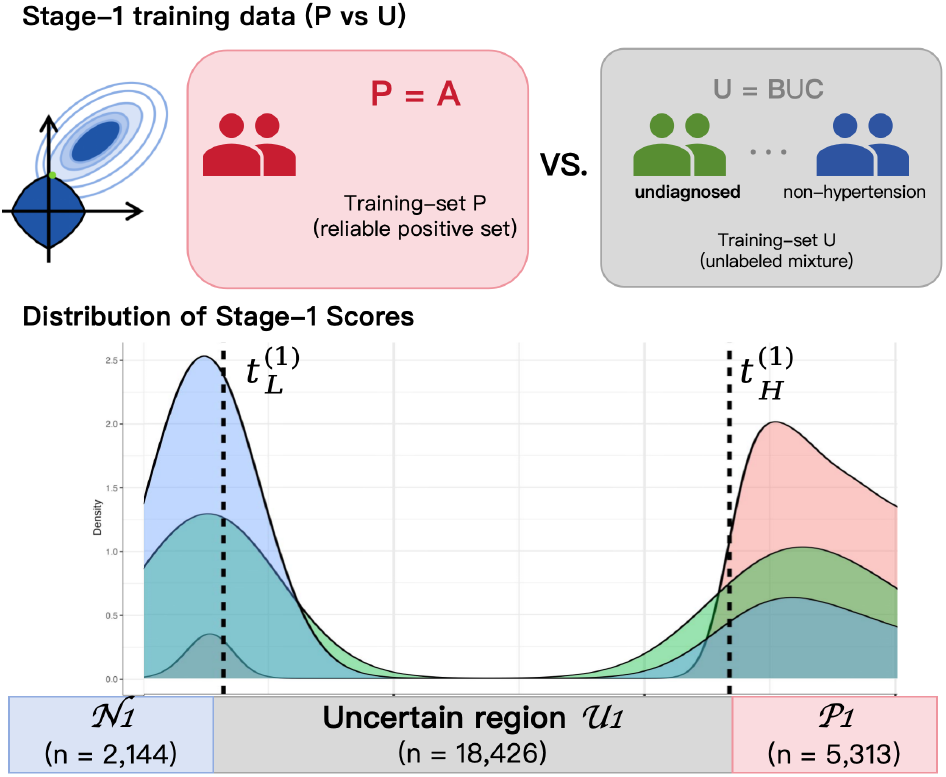
Stage-1 reliable-sample reconstruction using Elastic Net. Elastic Net logistic regression was fitted using only training-visible P/U labels to generate the stage-1 discrimination score *q*_*i*_ . Training-internal lower and upper thresholds 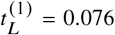 and *t* (1) = 0.598) 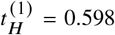 partitioned the training sample into candidate negatives *N*1 (n=2,144), the uncertain set *U*_1_ (n=18,426), and candidate positives *P*_1_ (n=5,313). Reference-group score distributions, when shown, are provided only for post hoc interpretation and were not used to determine the thresholds.

The stage-1 thresholds were selected empirically from the distribution of model-generated scores in the training data. The score distribution was inspected to identify lower- and upper-score regions representing relatively high-confidence candidate negatives and positives, while the broad intermediate region was retained as uncertain for further refinement. Based on this training-stage score distribution, the lower and upper thresholds were fixed at

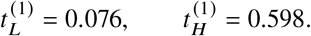

Accordingly, observations with *q*_*i*_ ≤ 0.076 were assigned to the stage-1 candidate-negative set *N*_1_, observations with 0.076 *< q*_*i*_ *<* 0.598 were retained in the uncertain set *U*_1_, and observations with *q*_*i*_ ≥ 0.598 were assigned to the stage-1 candidate-positive set *P*_1_. Threshold selection used only training-visible model scores and did not use the reference identities of groups B and C. Reference-group score distributions were examined only post hoc for interpretation of the selected score regions.

#### Stage 2: Nonlinear refinement of the uncertain region using random forests

Stage 2 is applied only to *U*_1_ to capture nonlinear relationships and interactions that are not adequately represented by the stage-1 linear ranking. Let

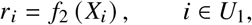

denote the second-stage ranking score produced by the random forest(Breiman, 2001; Liaw & Wiener, 2002). Let the stage-2 high- and low-confidence thresholds be 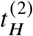 and 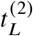, respectively, with 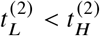 Then

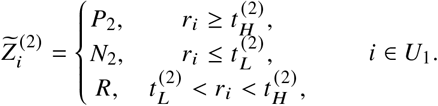

Here, *P*_2_ and *N*_2_ denote the stage-2 candidate-positive and candidate-negative sets, respectively, and *R* denotes the reject set containing observations that remain insufficiently certain after both stages. Observations in *R* do not contribute to the supervised XGBoost loss(Hendrickx et al., 2024).

The stage-2 random forest *f*_2_ was trained on the supervised set formed by the high-confidence candidates retained from stage 1. The stage-1 candidate positives *P*_1_ and candidate negatives *N*_1_ jointly define *D*_1_ and are assigned binary pseudo-labels 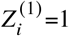and 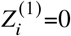 respectively:

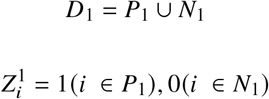

The fitted random forest *f*_2_ was then used to generate stage-2 scores *r*_*i*_ for observations in the stage-1 uncertain set *U*_1_. The distribution of these training-stage scores was subsequently examined to identify lower- and upper-score regions with greater classification confidence and a central region in which substantial uncertainty remained. Based on this distribution, the stage-2 lower and upper thresholds were fixed at approximately

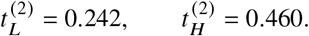

Observations with *r*_*i*_ *<* 0.242 were assigned to the stage-2 candidate-negative set *N*_2_, observations with 0.242 ≤ *r*_*i*_ *<* 0.460 were assigned to the reject set *R*, and observations with *r*_*i*_ ≥ 0.460 were assigned to the stage-2 candidate-positive set *P*_2_. As in stage 1, threshold selection was based only on training-stage model scores and did not use the reference identities of groups B and C. Reference-group distributions were used only for post hoc interpretation.

#### Final pseudo-labeled training set and class balancing

Candidate samples retained across the two stages were combined as

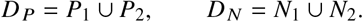

The final pseudo-label was defined as

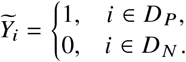

The final reconstructed training set contained 8,017 pseudo-positive and 8,017 pseudo-negative observations, for a total of 16,034 participants. Because this balancing step changes the training class prior, the final model output is interpreted primarily as a ranking and classification score rather than as an absolute population disease probability unless additional probability calibration is performed.

**Figure 4.**
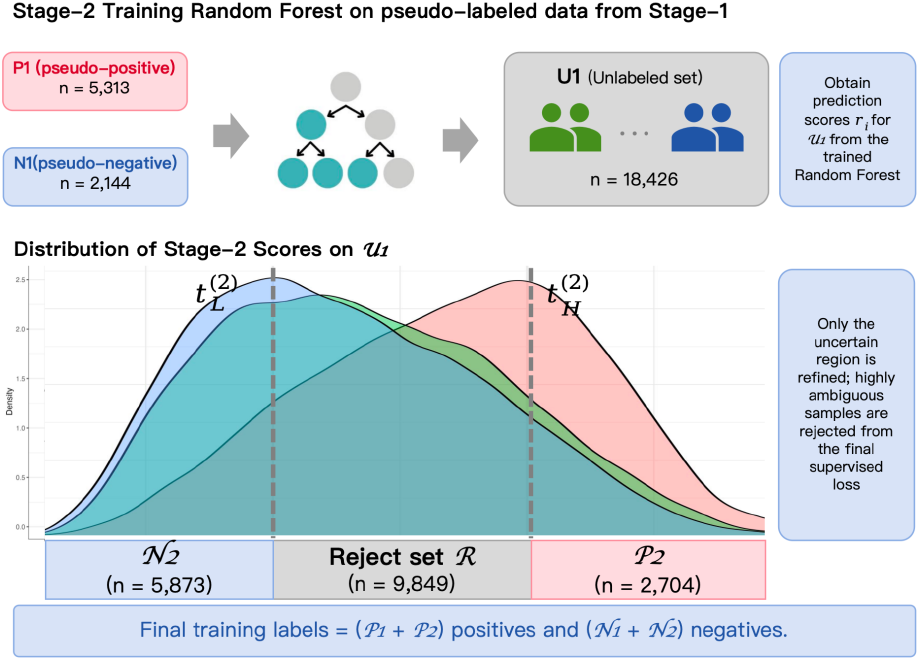
Stage-2 nonlinear refinement and uncertainty rejection. A random forest was trained using stage-1 candidate positives *P*_1_ and candidate negatives *N*_1_ as pseudo-labeled supervision and applied only to the uncertain set *U*_1_. The resulting score *r*_*i*_ and training-internal thresholds 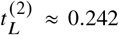 and 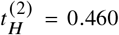 partitioned *U*1into candidate negatives *N*2 (n=5,873), the reject set R (n=9,849), and candidate positives *P*2 (n=2,704). Observations in R were excluded from the final supervised loss. Reference B/C identities were not used for model fitting or threshold construction.

### 2.8 Final classifier: XGBoost

After the two-stage reconstruction, XGBoost is trained on

{*X*_*i*_, 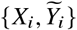 } to learn the final nonlinear decision function *g*(*X*).

The recorded final hyperparameters were

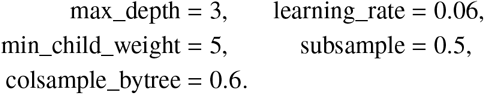

Hyperparameters were selected by grid search using AUC with respect to the reference disease status *Y* in the validation set as the model-selection criterion(Chen & Guestrin, 2016); the validation set was not used in the two-stage training-sample reconstruction. The final model produces a continuous output *P*_*i*_ for each participant. In the primary analysis, no additional probability calibration was applied and the classification threshold was fixed at 0.5:

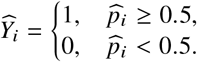

### 2.9 Baseline models

To evaluate the overall effect of two-stage training-set reconstruction, logistic regression, standard XGBoost, and a single-hidden-layer neural network were used as baseline models. The baseline models used the same predictors and data partition as the primary model but were trained directly on the original observable label *S* and evaluated against the reference disease status *Y* under the same validation/test framework. All models were evaluated in the same test set using a primary classification threshold of 0.5.

### 2.10 Model performance evaluation

Model performance was evaluated against the test-set reference disease status *Y* using the confusion matrix, accuracy, sensitivity, specificity, precision, negative predictive value, F1 score, and balanced accuracy. Because the intended use is screening rather than confirmatory diagnosis, particular emphasis was placed on sensitivity, the number of false negatives, and the sensitivity-specificity trade-off. Sensitivity and specificity were defined as

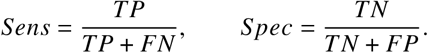

Balanced accuracy was defined as

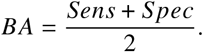

The relative reduction in false negatives compared with a baseline model was defined as

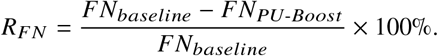

Model comparisons were descriptive. Because no paired resampling or other formal inferential procedure was used to compare model performance, numerical differences were not interpreted as statistically significant.

### 2.11 Model interpretation

SHAP (SHapley Additive exPlanations) values were used to attribute the final XGBoost output to individual input features(Lundberg & Lee, 2017). SHAP summary plots were used to compare global feature contributions, and dependence plots were used to display the direction and potential nonlinearity of model responses for selected variables. These analyses were used to interpret the predictive structure of the model and were not interpreted as estimates of causal effects.

### 2.12 Exploratory region-specific prevalence decision adjustment

To examine whether external region-specific hypertension prevalence information altered the classification operating point, two exploratory decision-adjustment strategies were evaluated. Let 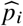 denote the original PU-Boost output, *r* (*i*) denote the region of participant *i, π*_*r* (*i*)_ denote the external hypertension prevalence for that region, and *π* denote the overall reference prevalence(Saerens et al., 2002; Wang et al., 2018).

The first strategy adjusted model output on the logit scale:

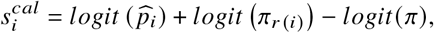

Where

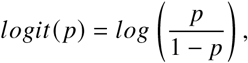

The adjusted probability was then computed as

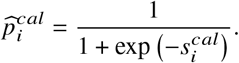

The second strategy directly used regional prevalence as a region-specific classification threshold:

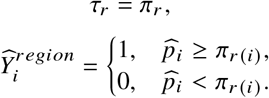

Both adjustments were evaluated in the same test set used for the primary analysis. They were intended to characterize how external prevalence priors shift the sensitivity-specificity operating point, rather than to demonstrate an improvement in the intrinsic discriminative ability of the model.

### 2.13 Software and reproducibility

All data processing, statistical analyses, model fitting, performance evaluation, and visualization were conducted in R version 4.6.0 (R Core Team, 2025). Elastic Net logistic regression was implemented with the glmnet package (Friedman et al., 2010), random forests with randomForest (Liaw & Wiener, 2002), gradient boosting with xgboost (Chen & Guestrin, 2016), and the single-hidden-layer neural network with nnet. ROC/AUC calculations were performed using pROC. SHAP values for the final XGBoost model were computed with fastshap and summarized and visualized with shapviz (Greenwell, 2024). Random seeds were fixed for cross-validation, random-forest fitting, negative-class downsampling, and other stochastic procedures to improve reproducibility.

## 3 Results

### 3.1 Cohort composition and label structure

A total of 43,105 participants were included. Based on prior diagnosis and reference disease status, participants were retrospectively classified into three groups: diagnosed hypertension (group A; n=13,287, 30.8%), undiagnosed hypertension (group B; n=9,393, 21.8%), and non-hypertensive individuals (group C; n=20,425, 47.4%). The A/B/C grouping was used to describe the reference disease structure of the cohort and for downstream model selection and performance evaluation. During two-stage PU reconstruction, only group A served as the labeled-positive set P; the reference identities of groups B and C were hidden from the training algorithm and together formed the unlabeled set U.

The data were split in a 6:2:2 ratio into training, validation, and test sets containing 25,883, 8,611, and 8,611 participants, respectively. The test set contained 3,987 reference-positive and 4,624 reference-negative participants. Model fitting preserved the P/U information structure, while the reference disease status was used only in the corresponding modelselection and performance-evaluation stages.

### Distributional overlap and label uncertainty within the unlabeled population

Exploratory group-level analyses indicated distributional differences among groups A, B, and C across multiple demographic, behavioral, medical-history, and anthropometric variables. However, these group-level differences did not translate into clear individual-level separation. Low-dimensional representations and score distributions showed substantial overlap between group B and group C in the available feature space.

These findings indicate substantial sample-level uncertainty between reference-positive and reference-negative observations within U, making the two components difficult to separate reliably using a single simple decision boundary. Accordingly, subsequent modeling focused on progressively identifying observations with higher label confidence while preventing highly uncertain observations from being forced into the final super-vised training set.

**Table 1.** Reference-group composition and label structure of the analytic cohort. Group membership was defined retrospectively from prior hypertension diagnosis and reference blood-pressure status. During PU reconstruction, group A constituted the labeled-positive set P, whereas groups B and C jointly constituted the unlabeled set U. Percentages are based on the full analytic cohort (N=43,105).

| Reference group | Clinical definition | n | % | Y | S | PU role |
| --- | --- | --- | --- | --- | --- | --- |
| A | Diagnosed hypertension | 13,287 | 30.8 | 1 | 1 | Labeled positive P |
| B | Undiagnosed hypertension | 9,393 | 21.8 | 1 | 0 | Unlabeled U |
| C | Non-hypertensive | 20,425 | 47.4 | 0 | 0 | Unlabeled U |
| Total | — | 43,105 | 100.0 | — | — | — |

**Figure 5.**
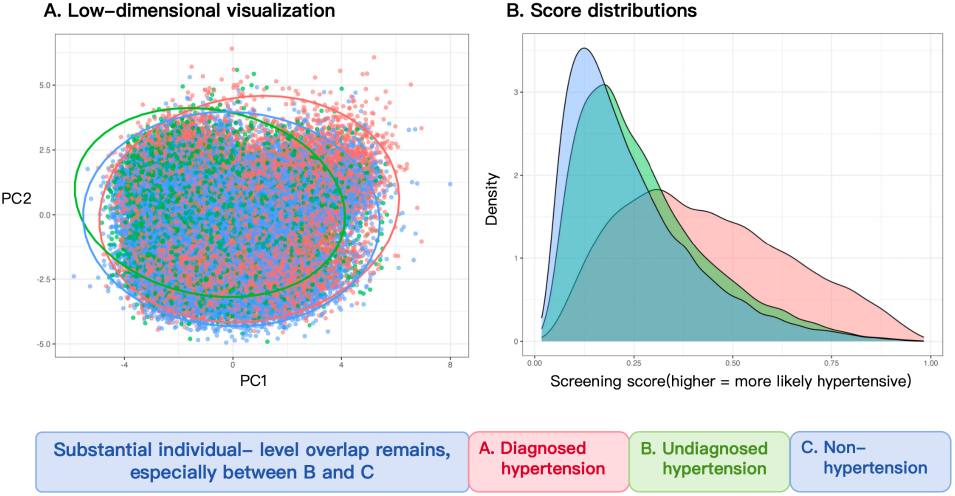
Distributional overlap among reference groups in the available predictor space. (A) Principal-component representation of diagnosed hypertension (A), undiagnosed hypertension (B), and non-hypertensive participants (C). (B) Distribution of the stage-1 P-versus-U score stratified retrospectively by reference group. Substantial individual-level overlap remained, particularly between groups B and C. Reference group identities are displayed for post hoc interpretation only and were not used during PU reconstruction.

### 3.2 Two-stage pseudo-label reconstruction

The training set was first scored by Elastic Net logistic regression for global P-versus-U stratification. Using the empirically selected training-stage thresholds of 0.076 and 0.598, the 25,883 training observations were partitioned into 5,313 candidate positives, 2,144 candidate negatives, and 18,426 observations in the uncertain set *U*_1_. Thus, 18,426 participants (71.2%) were retained in *U*_1_ for stage-2 nonlinear refinement.

Stage 2 applied a random forest only to *U*_1_. Using the stage-2 training-score thresholds of approximately 0.242 and 0.460, the 18,426 uncertain observations were further partitioned into 2,704 stage-2 candidate positives, 5,873 candidate negatives, and 9,849 highly ambiguous observations. The highly ambiguous subset was excluded from final classifier training, implementing a reject option for observations that remained insufficiently certain after both stages.

After combining candidates retained across both stages, the reconstructed training set contained 8,017 pseudo-positive and 8,017 pseudo-negative observations, for a total of 16,034 participants (61.9% of the original training set); the remaining 9,849 highly uncertain observations (38.1%) were excluded. The resulting class-balanced training sample was therefore composed of candidates retained at high confidence across the two reconstruction stages, while highly uncertain observations did not contribute directly to the final empirical loss.

### 3.3 Predictive performance comparison

PU-Boost was compared with three conventional supervised-learning models: logistic regression, standard XGBoost, and a neural network. At the prespecified classification threshold of 0.5, PU-Boost achieved a sensitivity of 0.8172 in the test set, the highest observed value among the compared models, versus 0.6832 for logistic regression, 0.6897 for standard XGBoost, and 0.7186 for the neural network. Relative to standard XGBoost, sensitivity increased by 0.1275 in absolute terms.

PU-Boost achieved a balanced accuracy of 0.6823, also the highest observed value, compared with 0.6624 for logistic regression, 0.6618 for standard XGBoost, and 0.6382 for the neural network. Overall accuracy was 0.6723 and likewise exceeded that of the three baselines. In contrast, the specificity of PU-Boost was 0.5474, lower than that of logistic regression and standard XGBoost.

The principal performance advantage of PU-Boost was therefore higher positive-case detection and balanced accuracy rather than uniform improvement across all classification metrics. Its test-set behavior reflected a high-sensitivity operating point, with improved identification of reference-positive cases at the cost of lower specificity.

### 3.4 Reduction in missed cases and screening-oriented performance

PU-Boost produced 729 false negatives in the test set, the fewest among the compared models. Logistic regression, standard XG-Boost, and the neural network produced 1,263, 1,237, and 1,122 false negatives, respectively. The corresponding reductions in false negatives with PU-Boost were 42.3%, 41.1%, and 35.0%. Compared with standard XGBoost, PU-Boost reduced the number of false negatives from 1,237 to 729, corresponding to 508 additional reference-positive participants being identified. This gain was accompanied by more false positives and lower specificity, indicating that the principal change was a redistribution of error types rather than a cost-free improvement in all aspects of classification performance.

This operating point is aligned with a screening objective that prioritizes reduction of missed cases: reference-positive individuals are more likely to be flagged for followup, while false-positive high-risk classifications can subsequently be resolved by standard blood-pressure measurement. PU-Boost is therefore better positioned as a high-sensitivity risk-stratification tool before confirmatory measurement than as a substitute for definitive clinical diagnosis.

**Figure 6.**
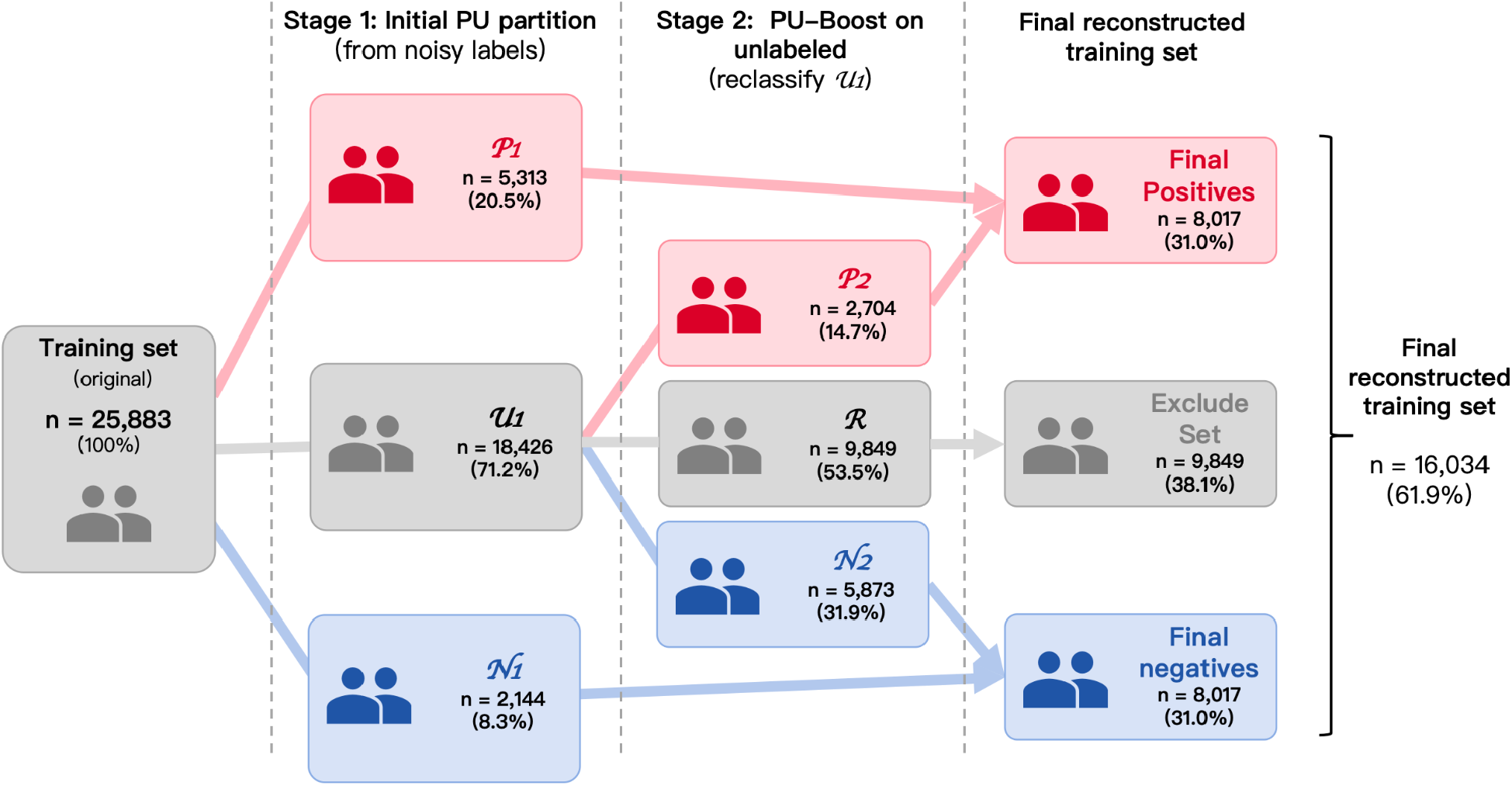
Sample flow through the two-stage reliable-sample reconstruction procedure. The original training set (n=25,883) was partitioned at stage 1 into *P*_1_ (n=5,313), *N*_1_ (n=2,144), and *U*_1_ (n=18,426). Stage 2 further divided *U*_1_ into *P*_2_ (n=2,704), *N*_2_ (n=5,873), and the reject set R (n=9,849). Combining retained candidates yielded 8,017 pseudo-positive and 8,017 pseudo-negative observations, forming a final reconstructed training set of 16,034 participants (61.9% of the original training set); 9,849 observations (38.1%) were rejected.

**Table 2.** Test-set predictive performance of PU-Boost and baseline models. All models were evaluated against the reference disease status Y in the same independent test set (n=8,611) using a classification threshold of 0.5. TP, true positives; TN, true negatives; FP, false positives; FN, false negatives; NPV, negative predictive value. Boldface indicates the highest observed value for each performance metric. Model comparisons are descriptive.

| Model | TP | TN | FP | FN | Accuracy | Sensitivity | Specificity | Precision | NPV | F1 | Balanced Accuracy |
| --- | --- | --- | --- | --- | --- | --- | --- | --- | --- | --- | --- |
| Logistic Regression | 2724 | 2967 | 1657 | 1263 | 0.6609 | 0.6832 | **0.6417** | **0.6218** | 0.7014 | 0.6511 | 0.6624 |
| XGBoost | 2750 | 2931 | 1693 | 1237 | 0.6597 | 0.6897 | 0.6339 | 0.6190 | 0.7032 | 0.6524 | 0.6618 |
| Neural Network | 2865 | 2579 | 2045 | 1122 | 0.6322 | 0.7186 | 0.5577 | 0.5835 | 0.6968 | 0.6440 | 0.6382 |
| PU Boost | 3258 | 2531 | 2093 | 729 | **0.6723** | **0.8172** | 0.5474 | 0.6089 | **0.7764** | **0.6978** | **0.6823** |

**Figure 7.**
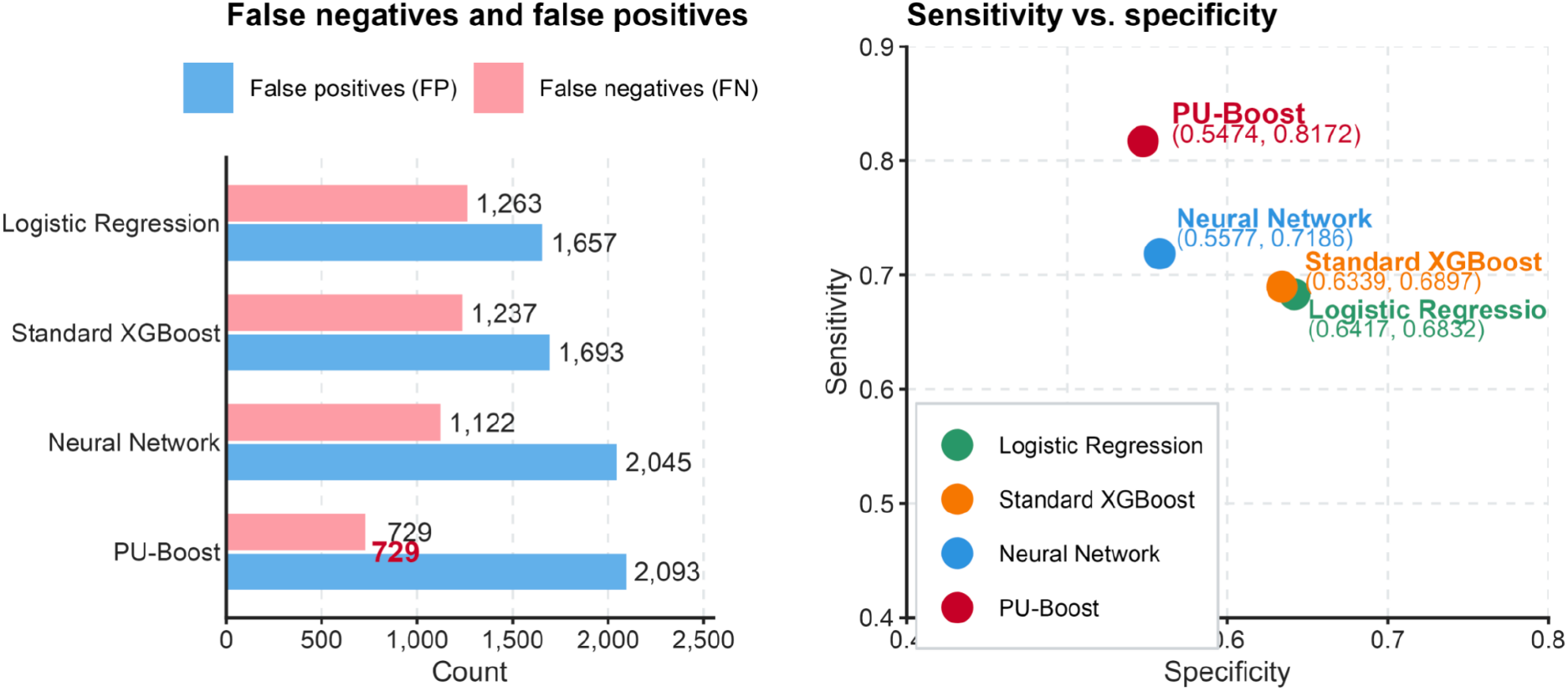
Test-set predictive performance and error profiles of PU-Boost and baseline models. (A) Sensitivity-specificity operating points at the prespecified classification threshold of 0.5. (B) Numbers of false negatives and false positives for logistic regression, standard XGBoost, the neural network, and PU-Boost. PU-Boost achieved the highest observed sensitivity and the fewest false negatives, accompanied by a higher number of false positives and lower specificity.

**Figure 8.**
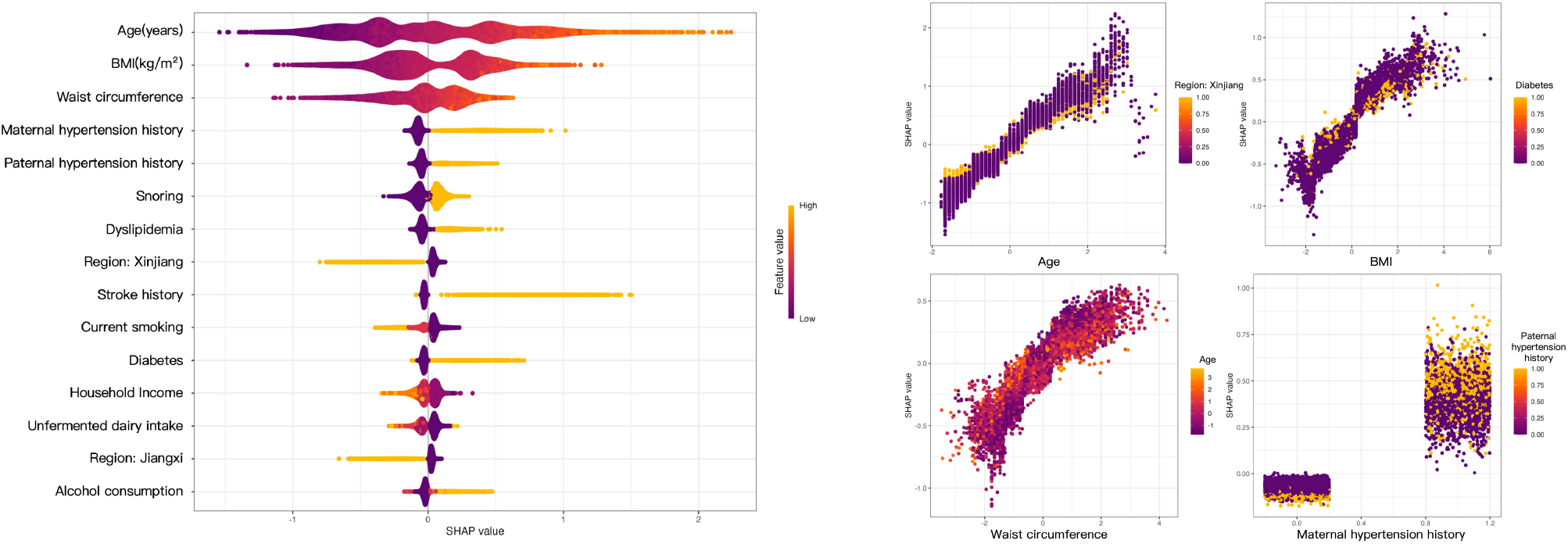
SHAP-based interpretation of the final PU-Boost model. (A) SHAP beeswarm plot showing the leading global contributors to model output, ordered by mean absolute SHAP value. Higher and lower feature values are indicated by the color scale. (B–E) SHAP dependence plots for age, body mass index, waist circumference, and maternal history of hypertension; color denotes the indicated secondary feature. SHAP values represent model attribution and should not be interpreted as causal effects.

### 3.5 Feature-importance analysis

SHAP analysis was used to quantify the contribution of each input feature to the final PU-Boost output. Age had the largest global SHAP contribution, followed by body mass index (BMI) and waist circumference. Other features among the ten highest-ranked global contributors included maternal history of hypertension, paternal history of hypertension, snoring, dyslipidemia, history of stroke, current smoking, and diabetes.

These highly ranked features are broadly consistent with established epidemiologic knowledge of hypertension and include age, general and central adiposity, family history, sleep-related factors, and metabolic and cardiovascular history. This concordance suggests that the final model relied substantially on clinically interpretable predictors, although SHAP values describe model attribution and do not establish causal effects or pathophysiologic mechanisms.

In addition to conventional risk-related variables, indicators for Xinjiang, Jiangxi, and Guangdong showed relatively large SHAP contributions, indicating that geographic information contributed to prediction. These contributions may reflect regional differences in exposure profiles, lifestyle, socioeconomic conditions, or access to health care, but the underlying mechanisms cannot be determined from SHAP analysis alone.

Overall, the major predictive contributions in PU-Boost were concentrated in variables with recognizable clinical and epidemiologic interpretations, providing feature-level insight into the model output.

### 3.6 Exploratory region-specific prevalence decision adjustment

As an exploratory analysis, we examined whether external region-specific hypertension prevalence information altered the operating point of PU-Boost. The unadjusted PU-Boost model had a sensitivity of 0.817, specificity of 0.547, F1 score of 0.698, and balanced accuracy of 0.682.

The region-logit adjustment increased specificity to 0.885 but reduced sensitivity to 0.369; the corresponding F1 score and balanced accuracy were 0.491 and 0.627. The region-specific prevalence-threshold adjustment produced similar results, with specificity of 0.871, sensitivity of 0.398, F1 score of 0.514, and balanced accuracy of 0.634.

**Figure 9.**
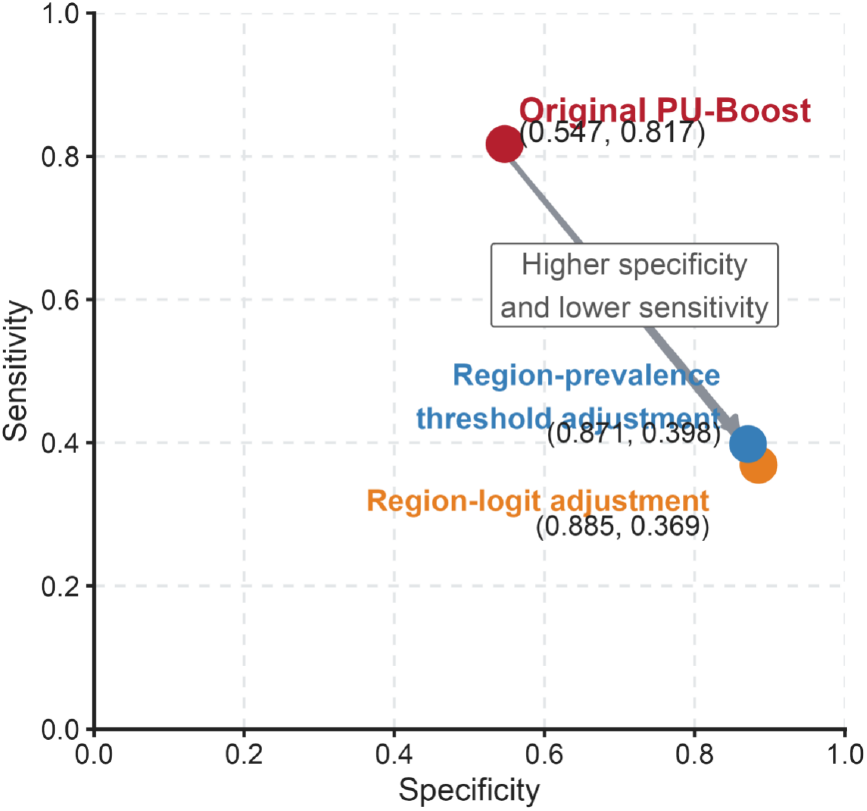
Sensitivity-specificity operating points under exploratory region-specific decision adjustments. The original PU-Boost operating point is compared with region-logit and region-prevalence threshold adjustments in the same test set. Both regional adjustments shifted the operating point toward higher specificity and substantially lower sensitivity, indicating a change in the decision rule rather than an improvement in intrinsic model discrimination.

Relative to the original operating point, the region-logit adjustment increased specificity by 0.338 while reducing sensitivity by 0.448. The prevalence-threshold adjustment increased specificity by 0.324 while reducing sensitivity by 0.419. Neither strategy improved the F1 score or balanced accuracy.

Thus, external regional prevalence information primarily shifted the sensitivity-specificity trade-off rather than improving overall screening performance under the metrics considered here. For a screening setting in which reduction of false negatives is prioritized, the original PU-Boost operating point retained substantially higher positive-case detection.

## 4 Discussion

This study formulates screening for undiagnosed hypertension as a positive-unlabeled learning problem and introduces PU-Boost, a two-stage reliable-sample reconstruction framework. The central issue is the mismatch between the observable diagnosis label and the reference disease status: training observes only labeled positives *P* = *A* and the unlabeled set *U* = *B* ∪ *C*, while U contains both hidden reference-positive and reference-negative individuals. A supervised classifier trained directly on S minimizes empirical risk with respect to the observable label, whereas the screening target is defined by Y(Bekker & Davis, 2020; Elkan & Noto, 2008); probabilistically, the corresponding targets are Pr(S=1|X) and Pr(Y=1|X). The methodological motivation for PU-Boost is therefore not to increase classifier complexity per se, but to reconstruct the training supervision so that hidden positives do not systematically contribute negative labels to the final classifier.

**Table 3.** Test-set performance under the original and region-specific decision rules. The original PU-Boost operating point was compared with region-logit and region-prevalence threshold adjustments using the same test-set predictions. Regional adjustments were exploratory and were intended to modify the sensitivity-specificity operating point rather than to improve the intrinsic discrimination of the fitted model.

| Method | Sensitivity | Specificity | Precision | F1 | Balanced Accuracy |
| --- | --- | --- | --- | --- | --- |
| PU Boost | **0.817** | 0.547 | 0.609 | 0.698 | **0.682** |
| Region-logit adjustment | 0.369 | **0.885** | 0.735 | 0.491 | 0.627 |
| Region-prevalence threshold adjustment | 0.398 | **0.871** | 0.727 | 0.514 | 0.634 |

The two-stage architecture of PU-Boost can be interpreted as regularized global ranking, local nonlinear refinement, and rejection of persistently uncertain observations(Bekker & Davis, 2020; Hendrickx et al., 2024; Liu et al., 2002). Elastic Net extracts stable global discrimination from the P/U label structure and separates readily classifiable observations from an uncertain region. A random forest is then applied only to this uncertain region to exploit nonlinear relationships and interactions. Observations that remain insufficiently certain after both stages are not forced into binary pseudo-labels but are excluded from final classifier training. The procedure therefore trades sample size for reduced direct contribution of highly uncertain supervision to the final empirical loss. Its potential benefit should be attributed to the joint reconstruction of the training distribution and supervision rather than to a simple sequence of Elastic Net, random forest, and XGBoost classifiers.

The independent test-set results were consistent with the intended screening behavior of this reconstruction strategy. At the prespecified threshold, PU-Boost achieved a sensitivity of 0.8172 and balanced accuracy of 0.6823, both the highest observed values among the compared models. Relative to standard XGBoost, sensitivity increased by 0.1275 and false negatives decreased from 1,237 to 729, a reduction of 508 cases (41.1%). These findings indicate that, for the same prediction task, PU-Boost tended to recover more reference-positive individuals missed by the conventional supervised baseline. However, the results support the overall two-stage reconstruction pipeline and do not establish that the performance difference arose solely from more accurate pseudo-labels. PU-Boost simultaneously modifies sample selection, the training class ratio, and the final operating point; without component-specific ablation analyses, their individual contributions cannot be separately quantified.

The most prominent change in error structure was a shift from false negatives toward false positives. PU-Boost produced fewer false negatives than the baseline models but had a specificity of 0.5474 and generated more false positives. Thus, the method did not improve every evaluation dimension simultaneously; rather, it operated at a more sensitivity-oriented screening point. From a decision perspective, this operating point is consistent with a screening context in which false negatives carry a higher relative cost. For hypertension screening, a false negative corresponds to a participant with reference hypertension being classified as low risk and potentially remaining unidentified, whereas a false positive can be reassessed by standard blood-pressure measurement. When the cost of missing disease exceeds the cost of additional confirmation, this error allocation may have practical screening value. Conversely, the lower specificity would be a substantial limitation if the model were used for definitive diagnosis. PU-Boost should therefore be viewed as a pre-confirmatory risk-stratification tool rather than a replacement for clinical diagnosis.

SHAP analysis provided further insight into the predictive structure of the final model. Age, BMI, waist circumference, parental history of hypertension, snoring, dyslipidemia, diabetes, history of stroke, and smoking contributed strongly to model output and are broadly consistent with established epidemiologic knowledge of hypertension(Herzog et al., 2025; Kolifarhood et al., 2019; Oparil et al., 2018; Peppard, 2000; Su et al., 2024). Several regional indicators also had relatively large SHAP contributions, suggesting that geographic information contributed to prediction, potentially through differences in exposure patterns, lifestyle, socioeconomic conditions, or health-care access(Zhou et al., 2019). Because SHAP quantifies attribution to model output rather than causal effects, these findings support clinical interpretability of the predictive structure but do not establish pathophysiologic mechanisms or exclude reliance on proxy variables or noise.

The exploratory prevalence-based decision adjustments further demonstrate the dependence of classification performance on the selected operating rule. Both the region-logit and region-specific threshold adjustments increased specificity but substantially reduced sensitivity and lowered both F1 score and balanced accuracy. External prevalence information therefore shifted the sensitivity-specificity operating point without improving discrimination under the metrics used here(Saerens et al., 2002; Wang et al., 2018). In other words, the threshold is a deployment-level decision parameter rather than an intrinsic property of discrimination; the same continuous score can map to different false-negative/false-positive trade-offs under different thresholds. For the present screening objective, which prioritizes reduction of missed cases, the unadjusted operating point was more consistent with the intended application.

Several limitations should be considered. First, PU learning assumes that labeled positives are sufficiently reliable, yet the process determining whether a hypertensive individual has been previously diagnosed is unlikely to be random. The labeling mechanism Pr(S=1|Y=1,X) may vary with healthcare access, health awareness, geography, socioeconomic status, and other covariates, producing positive-selection bias(Chang et al., 2022; Jain et al., 2016; Mielniczuk & Wawrzeńczyk, 2025). Second, two-stage sample selection, rejection of uncertain observations, and 1:1 class balancing jointly alter the composition and class prior of the training set, so the observed increase in sensitivity cannot be attributed uniquely to any single component(du Plessis et al., 2015; du Plessis et al., 2014; Kiryo et al., 2017; Li et al., 2022; Wu et al., 2023). Third, to preserve a simple screening setting, laboratory measurements, medication information, and direct blood-pressure measurements were intentionally excluded from predictors. This improves feasibility of data collection but may limit the achievable predictive performance. Fourth, the relatively low specificity of PU-Boost may increase false-positive volume and confirmatory workload in large-scale deployment, and its net practical value will depend on the screening cost structure. Fifth, the study population comprised community-dwelling adults aged ≥45 years from eight study areas in China. The findings should not be directly extrapolated to younger populations or other settings, and external validation in additional datasets is needed to establish transportability.

Future work should prioritize three directions. First, the sensitivity, false-negative reduction, and specificity of PU-Boost should be evaluated in independent external datasets to assess the stability of the training-sample reconstruction strategy. Second, the framework should be compared systematically with alternative PU-learning, weak-supervision, and mixture-model approaches, with component-specific analyses used where feasible to separate the effects of sample selection, class balancing, and uncertainty rejection. Third, decision thresholds should be optimized with respect to the relative costs of missed cases, confirmatory blood-pressure measurement, and acceptable false-positive burden in the intended screening workflow. Only after low false-negative rates and manageable confirmation burden are demonstrated externally should broader application to other screening problems with a reliable-positive plus mixed-unlabeled structure be considered.

## Conclusion

We developed PU-Boost, a two-stage reliable-sample reconstruction positive-unlabeled learning framework for screening undiagnosed hypertension. Under a label structure consisting of reliable positives and a mixed unlabeled population, PU-Boost progressively identifies high-confidence candidates, rejects persistently uncertain observations, and reconstructs the supervision used to train the final classifier. This provides a statistically explicit approach to identifying hidden positives under incomplete diagnostic labeling.

In the independent test set, PU-Boost achieved the highest observed sensitivity and balanced accuracy among the compared models and substantially reduced the number of false negatives. This gain was accompanied by lower specificity, but the resulting error structure is aligned with a screening workflow that prioritizes reduction of missed cases followed by confirmation with standard blood-pressure measurement. The model’s highest-contributing features were broadly consistent with established hypertension risk factors, supporting clinically interpretable prediction. Overall, PU-Boost provides a feasible and potentially useful framework for screening undiagnosed hypertension under incomplete diagnostic labels and motivates further work on external validation, threshold optimization, and evaluation in real-world screening settings.

## Data Availability

All data produced in the present study are available upon reasonable request to the authors

https://github.com/LionelWu11282562/A-Two-Stage-Probability-Stratified-Method-for-Ambiguous-Dataset-Optimization

## Author contributions

H.W.: Conceptualization, Methodology, Formal analysis, Investigation, Data curation, Software, Validation, Visualization, Project administration, Writing - original draft, Writing - review & editing.

L.S.: Conceptualization, Methodology, Formal analysis, Validation, Writing - review & editing.

H.Y.: Methodology, Software, Formal analysis, Validation, Visualization, Writing - review & editing.

B.L.: Conceptualization, Methodology. Q.W.: Writing - original draft, Software.

Z.Z.: Methodology, Writing - review & editing.

L.G.: Resources, Data curation, Writing - review & editing.

## Acknowledgements

The authors thank Professor Hao Helen Zhang for her helpful methodological discussions and valuable feedback during the development of this work.

## Data availability

Data are available from the corresponding author upon reasonable request, subject to applicable data-use requirements.

## Code availability

The analysis code, model implementation, and scripts used to reproduce the main results of this study are publicly available at: https://github.com/LionelWu11282562/A-Two-Stage-Probability-Stratified-Method-for-Ambiguous-Dataset-Optimization.

